# Extracting patient reported cannabis use and reasons for use from electronic health records: a benchmarking study of large language models

**DOI:** 10.64898/2026.03.06.26347824

**Authors:** Yiyu Wang, Selen Bozkurt, Nathan Le, Aishwarya Alagappan, Cho-Yi Huang, Swati Rajwal, Ashley Lewis, Jiyeong Kim, Titilola Falasinnu

**Author notes:** Contributed equally to this work and share first authorship (Yiyu Wang, Selen Bozkurt). **Corresponding Author:** Titilola Falasinnu, PhD, Division of Immunology and Rheumatology, Stanford School of Medicine, CA.

## Abstract

**Key messages:** **What is already known on this topic**

Cannabis use is increasingly documented in EHR narrative text, but structured fields do not capture use status or symptom related motivations, limiting research on pain self-management strategies in autoimmune rheumatic diseases.

**What this study adds**

We developed a natural language processing pipeline using large language models to identify cannabis use status (4-class) and reasons for use (6-class), with best performance from fine-tuned GatorTron for status and an LLM for reasons.

**How this study might affect research, practice or policy**

This scalable approach can support real-world evidence studies on symptom management, medication use, and outcomes among ARD populations, and it provides a methodological template for extracting under-documented patient behaviors from narrative notes.

**Objective:** The primary objective is to develop and evaluate a scalable and reproducible natural language processing (NLP) approach using large language models (LLM) to identify cannabis use status and reasons for cannabis use among patients with autoimmune rheumatic diseases (ARDs) from unstructured electronic health record (EHR) clinical notes.

**Methods and Analysis:** We conducted a retrospective study using EHR clinical notes from patients with ARDs (2015–2024). Notes were screened for cannabis-related mentions using fuzzy string matching against a curated keyword lexicon with a similarity threshold of 90, extracting 50-word context windows (±25 words). Two domain experts annotated 886 randomly sampled snippets across four classes: (1) not a true cannabis mention/uncertain, (2) denial of use, (3) positive past use, and (4) positive current use. Using these annotations, we compared multiple LLM prompting strategies (zero-shot to few-shot; temperature tuning) and a fine-tuned clinical model (GatorTron 345M). For “reason for use,” 1,027 snippets were annotated into six categories: pain, nausea, sleep, anxiety/stress/mood, appetite, and not mentioned/unknown. Models were evaluated on a held-out validation set using accuracy, F1, recall, and precision. We then aggregated snippet-level predictions to patient level to describe temporal trends and subgroup differences.

**Results:** For cannabis use status classification, the fine-tuned GatorTron model achieved the highest performance (accuracy 0.90; F1 0.91; recall 0.90; precision 0.90). For the reason of cannabis use classification, GPT-OSS-20B achieved the highest performance (accuracy 0.90; F1 0.90; recall 0.90; precision 0.92). Patient-level analyses characterized trends in documented cannabis use from 2015–2024 and compared clinical characteristics between current users and patients denying use.

**Conclusion:** High-precision extraction of cannabis use status and reasons for use from EHR notes is feasible using a combination of fine-tuned clinical language models and LLM-based classifiers. This approach enables scalable measurement of patient-reported symptom self-management strategies in ARDs, supporting observational research and potential clinical decision support.

## Introduction

Autoimmune rheumatic diseases (ARDs) are chronic inflammatory conditions often accompanied by chronic pain, functional disability, and reduced quality of life(1). Chronic pain remains difficult to manage with existing treatments. Cannabis has emerged as a potential adjunct for pain through modulation of endocannabinoid receptors involved in pain processing(2–4). However, evidence on cannabis use in ARDs remains limited. As such, the National Pain Strategy calls for proof-of-concept analyses using large health care databases to identify pharmacological and non-pharmacological treatment patterns among people with chronic pain and related diagnostic clusters(5).

Electronic health records (EHR) offer opportunities to examine cannabis use in clinical populations(6–10), but structured fields rarely capture cannabis status, clinical context, or patient-reported reasons for use. These details are especially relevant in ARDs, where pain and other symptoms may motivate self-management outside prescribed therapy, and are often embedded in the unstructured narrative clinical notes, Recent advances in natural language processing (NLP) via large language models (LLMs) have enabled the analysis of unstructured clinical text without requiring manual data annotation(11). By leveraging large-scale pretraining, these models can represent contextual information in narrative text with reduced reliance on task-specific rule engineering(12,13). As a result, LLMs are increasingly used for automated phenotyping and information extraction from EHR data(6,14–16). Despite the growing use of LLMs, important questions regarding the reliability, generalizability, and behaviors of LLMs in clinical tasks remain under-investigated(17). Specifically, there are ongoing debates about the relative advantages of general-purpose versus clinically pretrained domain-specific models(18–20), as well as how design choices such as prompting strategies and decoding parameters influence performances(21).

This study addresses two gaps: (1) whether LLMs can accurately identify cannabis use and associated clinical contexts, including reasons for use, and (2) how performance varies across decoding temperatures and prompt strategies. We compare five models, including general-domain (GPT-OSS, LLaMA, Gemini) and clinical-domain models (GatorTron, MedGemma), using expert-annotated EHR snippets and standard NLP metrics. Our goal is to clarify the strengths, limitations, and practical use of LLM-based phenotyping for pain management and population health research.

## Methods

### Study design and data source

We conducted a retrospective observational study using EHR data from a tertiary academic medical center. The dataset comprised longitudinal EHR data, including unstructured clinical notes and structured demographic, diagnostic, medication, and encounter information, collected during routine clinical care between 2015 and 2024. The study was approved by the Stanford University Institutional Review Board (IRB #53750) with a waiver of informed consent for the secondary use of de-identified data. No patients or members of the public were involved in the design, conduct, reporting, or dissemination of this research.

### Study population

The study population included patients aged ≥18 years who received care at an outpatient rheumatology or joint immunology-dermatology clinic within a major academic healthcare institution in Northern California between 2015 and 2024. Patients were included if they had one or more ARDs, documented at least twice and recorded ≥3 months apart for ankylosing spondylitis (AS), psoriatic arthritis (PsA), rheumatoid arthritis (RA), Sjögren’s syndrome (SjS), systemic lupus erythematosus (SLE), or systemic sclerosis (SSc), based on ICD-10-CM codes reported.

### Model development data set

For model development, we created three non-overlapping subsets to prevent information leakage: training notes (n=791, including snippets used in LLM prompts), validation/testing notes (n=40, 10 per class), and held-out notes for final application at scale (n=42,858). Notes classified as current or past cannabis use were further selected for reason-of-use analysis (n=19,498), including annotated notes (n=1,027), validation data (n=60, 10 per class), and an application set (n=19,426; Figure 1, Figure S1). All partitions were defined before model training and evaluation.

**Figure 1.**
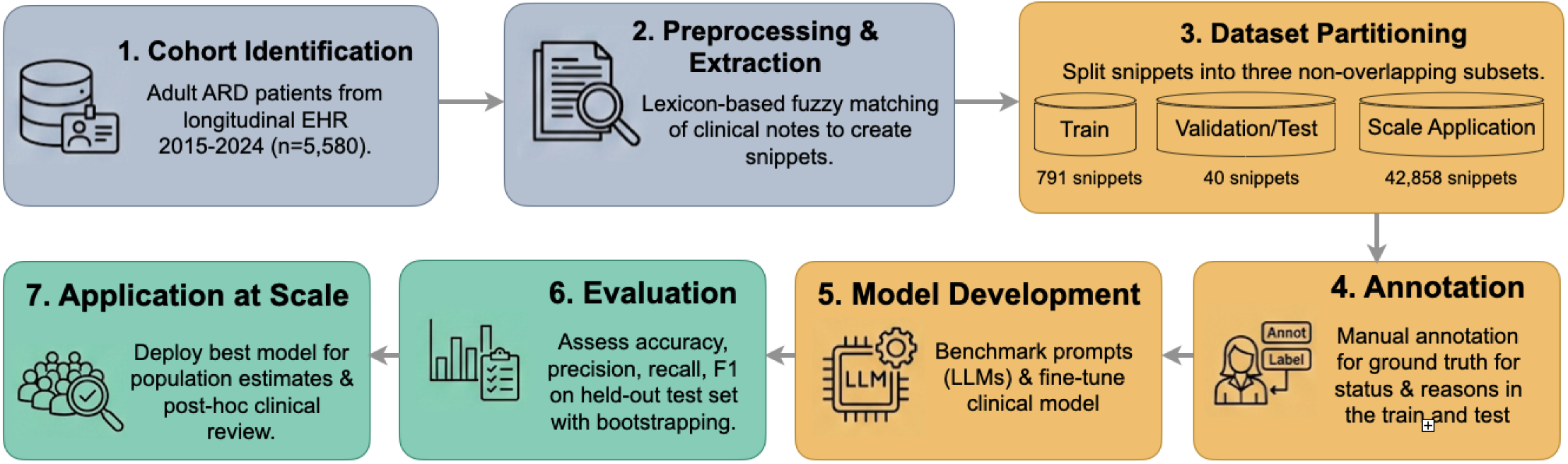
Study Design.

### Text preprocessing

We processed clinical notes to identify candidate snippets containing cannabis-related terms. We used an established lexicon containing cannabis keyword (9) select notes with at least one term. To capture misspellings, typographical errors, and lexical variation in clinical text, approximate string matching was performed with rapidfuzz(22).

For each matched term, a fixed-length context window of 50 words was extracted, consisting of 25 words preceding and 25 words following the matched term. Extracted snippets were normalized by removing non-English characters and punctuation, while retaining relevant symbols (“:”, “/”, “-”, “#”, “.”, and “,”) which commonly occur in semi-structured clinical notes (e.g., field-value pairs such as “Smoking: Yes” and medication or identifier notation). This process yielded a corpus of 43,689 cannabis-relevant snippets from 3,244 patients.

### Manual annotation and reference standard construction

We adapted annotation guidelines from prior work(7). Two domain experts independently annotated snippets over three iterative rounds to refine label definitions and resolve disagreements.

First, a random sample of 831 cannabis-relevant snippets was labeled into four mutually exclusive categories: (1) not a true cannabis mention/uncertain, (2) denial of use, (3) positive past use, and (4) positive current use. The class distribution was 28.41% not true/uncertain, 10.42% denial, 16.61% past use, and 44.56% current use.

Second, an independent sample of 1,027 snippets was annotated into six categories: pain, nausea, sleep, anxiety/stress/mood, appetite, and not mentioned/unknown. The resulting distribution was 61.44% not mentioned/unknown, followed by pain (15.38%), nausea (9.74%), sleep (7.50%), anxiety/stress/mood (4.58%), and appetite (1.36%), highlighting class imbalance and the frequent absence of explicit documentation of use motivations in clinical notes.

### Information extraction using large language models (LLM)

We evaluated five LLMs for cannabis use extraction from clinical notes, including both general-domain (Gemini-2.0, LLaMA-3.1, GPT-OSS-20B), and clinically pretrained models (MedGemma-4B, and GatorTron-345M). First, to assess the impact of supervised domain adaptation, we fine-tuned a clinical language model, GatorTron-345M, for cannabis use status classification(23). Fine-tuning was performed using manually annotated snippets that were not included in prompt development or validation. Learning rate, dropout, and maximum sequence length were optimized with Bayesian optimization using Optuna(24). Training data were split 80/20 for hyperparameter selection, and the configuration with the lowest validation loss was retained (Table S3). The fine-tuned model was evaluated on the same held-out validation set used for prompt-based models. Second, we developed and evaluated eight strategies (chain-of-thoughts, zero-shot, one-shot, two-shots, structured reasoning, confidence scoring, multi-step verification, uncertainty handling) for using LLMs for cannabis use extraction, informed by established best practices in the literature (Table S1, S2). Prompt selection and optimization were conducted using a stratified validation set consisting of 10 annotated snippets per class for each classification task.

We evaluated decoding temperatures ranging from 0.0 to 1.0, prioritizing lower temperatures to improve reproducibility and reduce output variability. We evaluated model performance using accuracy, precision, recall, and F1 score for both cannabis use status and reason-for-use classification. All calculations were performed on the held-out data not used for training or prompt development. Confidence intervals were estimated with 10,000 bootstrap resamples of the held-out dataset.

### Assessment of clinical utility

The best-performing models were applied to the remaining data (use status: n=42,858 and use reason: n=19,426) to generate population-level estimates of cannabis use status and reasons. Snippet-level predictions were aggregated to the patient level and summarized with frequency distributions, temporal trends, and subgroup comparisons by disease and demographic category.

In addition, these aggregate patterns including the distribution of reported reasons for use and longitudinal trends, were summarized and compared with domain expert expectations and findings from prior clinical and epidemiological literature.

## Results

The final study cohort included 5,580 adult patients with ARDs, contributing over 2 million clinical notes between 2015 and 2024. The demographic characteristics of the cohort included 4,286 (76.81%) females, 3,477 (62.31%) white patients, and a mean age of 59.75 years (at the most recent medical encounter).

Demographic and clinical characteristics of the study population are summarized in Table 1, including age, sex, race/ethnicity, disease type, insurance type, and baseline clinical diagnosis. Overall, the majority had rheumatoid arthritis, and the cohort was predominantly female and White, consistent with the underlying clinic population.

**Table 1.** Cohort Characteristics of Patients with Cannabis-matched Clinical Notes.

| Variables |  | M | SD | Range |
| --- | --- | --- | --- | --- |
| Age* | (years) | 60.9 | 19.1 | 17 - 100 |
|  |  | N | (%) |  |
| Gender | Female | 2516 | 77.6 |  |
|  | Male | 728 | 22.4 |  |
| Ethnicity | Non-Hispanic | 2632 | 81.1 |  |
|  | Hispanic | 449 | 13.8 |  |
|  | Unknown | 163 | 5.0 |  |
| Race | White/Caucasian | 2034 | 62.7 |  |
|  | Black/African | 106 | 3.3 |  |
|  | Asian | 525 | 16.2 |  |
|  | Native Hawaiian or Pacific Islander | 30 | 0.9 |  |
|  | American Indian or Alaska Native | 22 | 0.7 |  |
|  | Other or unknown | 527 | 16.2 |  |
| Marital Status | Married | 1884 | 58.1 |  |
|  | Single | 801 | 24.7 |  |
|  | Widowed | 289 | 8.9 |  |
|  | Divorced | 195 | 6.0 |  |
|  | Life Partner | 31 | 1.0 |  |
|  | Separated | 14 | 0.4 |  |
|  | Other | 30 | 0.9 |  |
| Insurance Type | Public | 1647 | 50.8 |  |
|  | Private | 1386 | 42.7 |  |
|  | Unknown | 211 | 6.5 |  |
| Disease Condition<sup>†</sup> | Ankylosing spondylitis (AS) | 188 | 4.5 |  |
|  | Psoriatic arthritis (PsA) | 494 | 11.9 |  |
|  | Rheumatoid arthritis (RA) | 1837 | 44.4 |  |
|  | Sjogren syndrome (SjS) | 612 | 14.8 |  |
|  | Systemic lupus erythematosus (SLE) | 596 | 14.4 |  |
|  | Systemic sclerosis (SSc) | 415 | 10.0 |  |

\*Age is calculated from the most recent encounter date - date of birth.
† The total exceeds 3,244 because 763 patients received multiple diagnoses.

### Extraction of cannabis use

Across prompting strategies and temperatures, GPT-OSS-20B had the strongest performance among generative models for cannabis use status classification (Figure 2, top row; Figure S2). No prompt strategy consistently outperformed the others, indicating model-dependent prompt sensitivity (Figure 2a). Performance generally improved at lower temperatures (Figure 2b), and each model’s best classification results are shown in Table 2.

**Figure 2.**
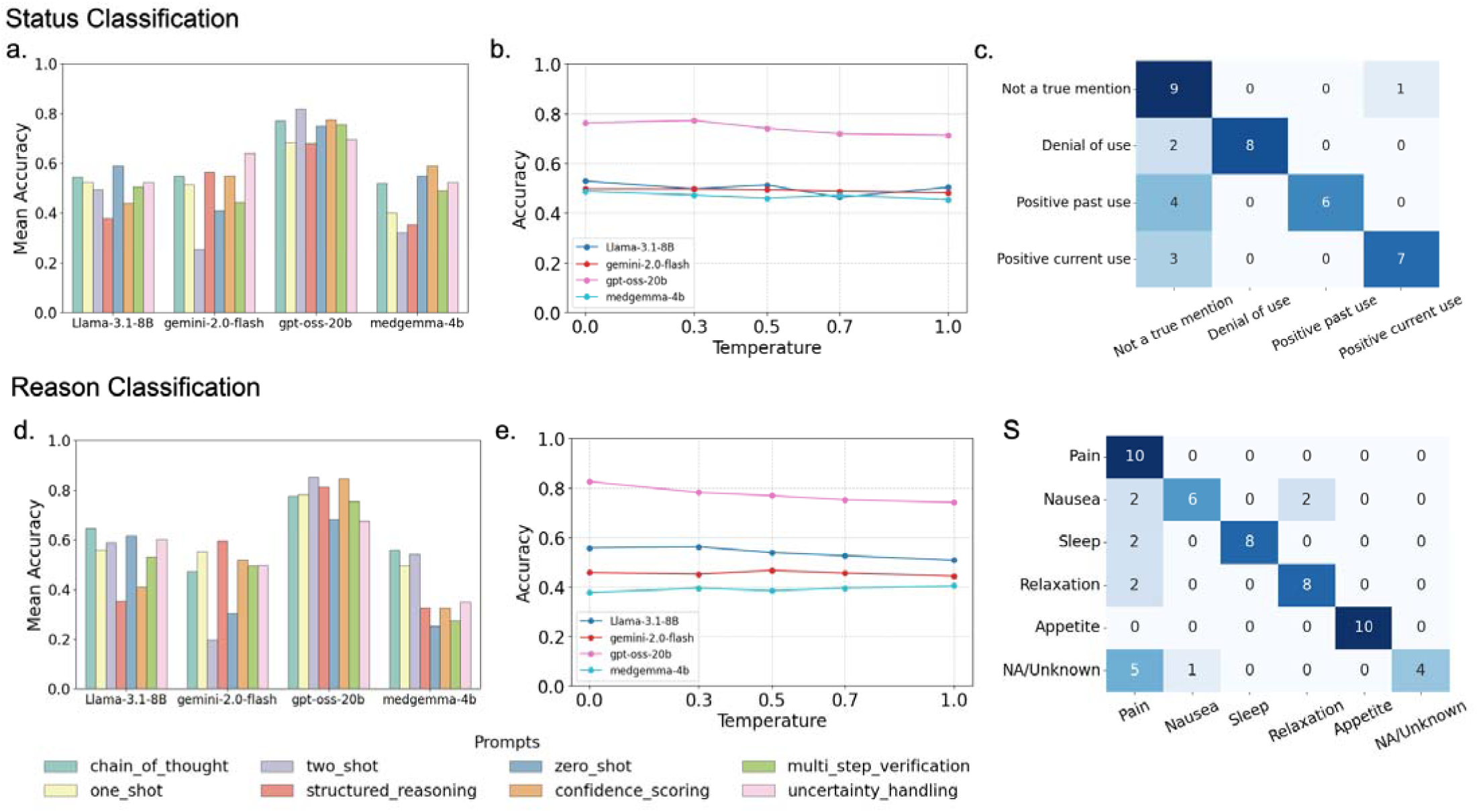
LLM performance for classifying cannabis use status (top row) and reasons for use (bottom row). Status classification: (a) Mean accuracy of each prompting strategy across decoding temperatures and models, showing that no single strategy consistently performs well across models. (b) Mean accuracy aggregated across prompting strategies by model and temperature. GPT-OSS-20B (pink) achieves the highest performance among generative LLMs but does not outperform the fine-tuned GatorTron model. Lower decoding temperatures are associated with slightly improved performance. (c) Confusion matrix for GPT-OSS-20B using a chain-of-thought prompt on the test set. Reason classification: (d) Mean accuracy of each prompting strategy across decoding temperatures and models, again demonstrating variability in prompt effectiveness across models. (e) Mean accuracy aggregated across prompting strategies by model and temperature, with GPT-OSS-20B (pink) again achieving the highest performance among generative LLMs. Lower decoding temperatures are associated with improved performance. (f) Confusion matrix for GPT-OSS-20B using a two-shot prompt at temperature 0.3 on the test set (10 samples per class).

**Table 2.** Best Performance Per Model in Classifying Cannabis Use Status and Reason.

| Task | Model | Strategy | Accuracy | F1 | Recall | Precision |
| --- | --- | --- | --- | --- | --- | --- |
| Status of Cannabis Use | **Gemini-2.0-flash** | Zero-shot | 0.70<br>(0.58, 0.83) | 0.69<br>(0.55, 0.82) | 0.70<br>(0.58, 0.83) | 0.72<br>(0.59, 0.85) |
|  | **MedGemma-4B** | Structured Reasoning | 0.65<br>(0.5, 0.78) | 0.64<br>(0.49, 0.78) | 0.65<br>(0.50, 0.78) | 0.65<br>(0.50, 0.80) |
|  | **GPT-OSS-20B** | Chain of thought | 0.85<br>(0.75, 0.95) | 0.83<br>(0.75, 0.95) | 0.85<br>(0.75, 0.95) | 0.88<br>(0.80, 0.96) |
|  | **Llama3.1-8B** | Structured reasoning | 0.63<br>(0.48, 0.78) | 0.63<br>(0.47, 0.77) | 0.63<br>(0.48, 0.77) | 0.68<br>(0.53, 0.82) |
|  | **GatorTron Fine tuned\*** | - | 0.90<br>(0.80, 0.98) | 0.90<br>(0.80, 0.97) | 0.90<br>(0.80, 0.98) | 0.91<br>(0.84, 0.97) |
| Reason for Cannabis Use | **Gemini-2.0-flash** | One-shot | 0.60<br>(0.50, 0.68) | 0.54<br>(0.44, 0.62) | 0.60<br>(0.50, 0.68) | 0.51<br>(0.41, 0.60) |
|  | **MedGemma-4B** | Multistep verification | 0.60<br>(0.48, 0.72) | 0.60<br>(0.47, 0.71) | 0.60<br>(0.48, 0.72) | 0.67<br>(0.57, 0.78) |
|  | **GPT-OSS-20B** | Two-shot | 0.90<br>(0.83, 0.97) | 0.90<br>(0.82, 0.97) | 0.90<br>(0.83, 0.97) | 0.92<br>(0.87, 0.97) |
|  | **Llama3.1-8B** | Structured reasoning | 0.72<br>(0.62, 0.80) | 0.69<br>(0.59, 0.78) | 0.72<br>(0.62, 0.80) | 0.80<br>(0.58, 0.86) |
|  | **GatorTron Fine-tuned\*** | - | 0.53<br>(0.43, 0.63) | 0.55<br>(0.39, 0.59) | 0.53<br>(0.43, 0.63) | 0.55<br>(0.45, 0.64) |

\* No prompt strategies or decoding temperatures were evaluated for GatorTron, as it was fine-tuned as an encoder-based classification model and does not perform text generation.
The lower and upper bound of the 95% confidence interval from the bootstrapping are presented in the parentheses.

Fine-tuned GatorTron achieved the highest overall performance on the held-out validation set (accuracy = 0.90, 95% CI [0.75, 0.96]; F1 = 0.91, 95% CI [0.75, 0.96]; Table 2). Performance was similar for current use (F1 = 0.90) and past use (F1 = 0.95).

### Extraction of reason of cannabis use

We next evaluated LLM performance on the more challenging task of classifying reasons for cannabis use from clinical notes. We repeated the same systematic benchmarking procedure across models, prompts, and temperatures (Figure 2, bottom row; see Figure S3 for detailed performance by temperature, prompts, and models).

GPT-OSS-20B performed best overall, achieving the highest average accuracy across prompts and temperatures and the best single configuration (accuracy = 0.90, 95% CI [0.69, 0.92]; F1 = 0.90, 95% CI [0.69, 0.91]; Table 2). As with status classification, no prompt strategy was consistently superior, and lower temperatures improved performance.

Based on these results, GPT-OSS-20B using a two-shot prompt strategy and a lower temperature decoding (temp=0.3) was selected to annotate cannabis-related reasons in all snippets labeled as current or past use. These results demonstrate that modern LLMs can be effectively leveraged to automatically extract not only cannabis use status but also contextual information about motivations and reasons for use.

### Generating Clinical Insights

To illustrate clinical utility, we applied the best-performing pipeline to annotate the rest of the notes from 2015 to 2024 (Figure 3a). Overall, 1,101 patients (19.7%) had at least one note indicating current cannabis use, although annual documentation remained approximately 10%. The proportion reporting cannabis use increased from 3% in 2015 to 10% in 2024 (Figure 3b), with similar upward patterns by sex and across ARD conditions (Figure 3c,d). We compared current users, defined as patients with at least one current-use note, with non-users whose notes documented denial of use. Charlson Comorbidity Index scores did not differ significantly, suggesting comparable comorbidity burden (Figure 3e), whereas current users had higher medication counts (Figure 3f). Interestingly, annual pain scores were higher among current users than non-users before 2023, but this pattern reversed in 2023, when non-users reported slightly higher mean pain scores (Figure 3g). While these associations do not establish temporal order or causality, they provide insights about the pain experiences of ARD patients, as pain remained the predominant LLM-extracted reason for cannabis use across the decade. Additionally, sleep-related reasons increased and became the second most frequent indication beginning in 2016 (Figure 3h), underscoring the need to address sleep disturbance as part of comprehensive ARD care.

**Figure 3.**
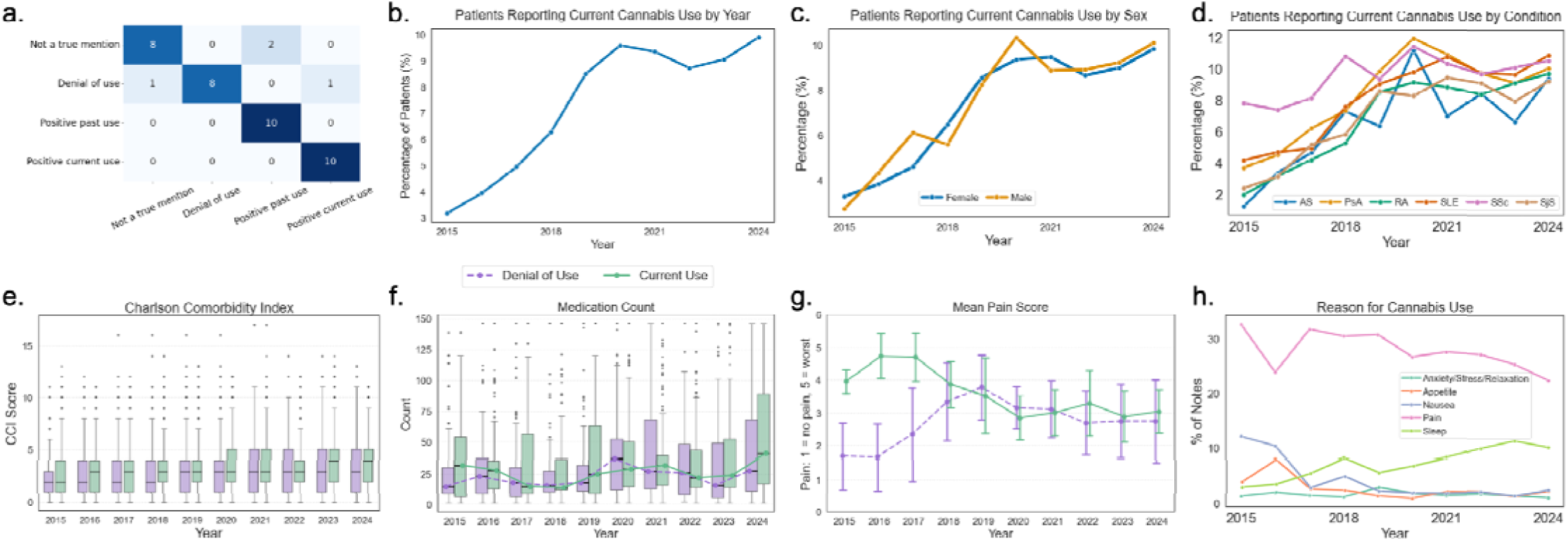
Clinical Insights from LLM applications. (a) Confusion matrix for cannabis use status classification using the fine-tuned GatorTron model. (b) Percentage of patients reporting current cannabis use over time. (c) Sex distribution among current cannabis users across years. (d) Distribution of current cannabis users across autoimmune rheumatic disease (ARD) conditions. (e) Charlson Comorbidity Index scores showing no significant differences between cannabis users and non-users. (f) Medication counts indicating higher overall medication use among cannabis users compared with non-users. (g) Longitudinal pain scores, showing higher reported pain among cannabis users prior to 2023, with non-users reporting higher pain scores in 2023–2024. (h) Trends in reported reasons for cannabis use over 10 years; pain remained the predominant indication, while sleep-related use (light green) became the second most frequently reported reason beginning in 2016.

### Computational performance

All models were run on two NVIDIA GeForce RTX 5090 GPUs (32 GB VRAM per GPU). Running GPT-OSS-20B required a peak GPU memory usage of approximately 21.9 GB per GPU, with an average processing time of ∼8.9 seconds per batch (batch size = 8 snippets).

## Discussion

In this study, we systematically evaluated the role of LLMs in extracting cannabis use phenotypes from unstructured EHR notes and compared prompt-based generative approaches with supervised domain-adapted transformers. Our findings demonstrate that model scale alone does not guarantee superior performance in clinical information extraction. Instead, performance advantages were task-dependent: for more structured classification tasks such as cannabis use status, a fine-tuned clinical transformer outperformed larger generative LLMs, whereas for the more contextually complex task of identifying reasons for cannabis use, generative LLMs demonstrated relative advantages.

Our study contributes to empirical characterization of LLM behavior in biomedical informatics, where mechanistic understanding remains limited(17,25). Prior work shows that LLM performance can be sensitive to prompts, especially in reasoning and multiple-choice tasks(26–28), while temperature effects vary by task and model size(21,29). Consistent with this literature, no prompt strategy generalized across models in our benchmark, supporting model-specific evaluation rather than assuming prompt transferability.

This study extends prior work on LLM-based EHR phenotyping in several ways. Other work on substance use extraction has compared generallZlpurpose LLMs with conventional neural and gradientlZlboosting approaches on annotated EHR corpora, highlighting task-dependent tradelZloffs in accuracy, efficiency, and implementation complexity(14,15). In contrast, our study focuses on a specific patient-reported behavior (i.e., cannabis use) and associated reasons for use in rheumatology, systematically comparing multiple general-domain and clinically pretrained LLMs with a fine-tuned clinical transformer on expert-annotated snippets from routine rheumatology notes. By jointly evaluating status classification and more semantically complex reason-for-use categorization, and then scaling these phenotypes to population-level trends over a decade, this work provides one of the first detailed characterizations of when smaller, domain-adapted models outperform large generative LLMs and when the latter add value for context-rich extraction in a real-world rheumatology setting

### Task-dependent value of LLMs

For cannabis use status classification, fine-tuned GatorTron achieved the strongest and most stable performance. This task involves relatively constrained categories (current, past, denial, not true mention) and explicit lexical cues, which may favor supervised domain adaptation over large generative reasoning models. These findings suggest that, for well-defined extraction tasks with annotated data available, smaller domain-specific transformers may offer greater efficiency and comparable or superior performance relative to computationally intensive generative LLMs.

In contrast, reason-for-use classification is inherently more semantically heterogeneous and context-dependent. Motivations for cannabis use are often embedded in narrative explanations, implicit statements, or co-occurring symptom descriptions. In this setting, GPT-OSS-20B outperformed other models, suggesting that generative LLMs may provide advantages for higher-entropy tasks requiring broader contextual reasoning.

Taken together, our results indicate that the benefits of LLMs in clinical NLP are not universal but depend on the linguistic complexity and structural constraints of the task.

### Computational cost and practical considerations

The computational requirements of large generative models were substantial, requiring high-memory GPUs and non-trivial inference time. Given that fine-tuned GatorTron achieved superior performance for use-status classification with far fewer parameters, our findings raise important practical considerations regarding the trade-off between computational cost and incremental performance gains in clinical pipelines. For healthcare systems seeking scalable deployment, domain adaptation of smaller models may be a more resource-efficient strategy for structured phenotyping tasks, while reserving large LLMs for tasks where contextual inference is essential.

### Generalizability and overfitting

Although fine-tuned GatorTron achieved high validation performance, strong in-domain validation performance might not translate to stable out-of-sample generalization in clinical NLP tasks. Explicit evaluation on independent annotated samples remains essential, particularly when deploying models for large-scale phenotyping. Similarly, we observed that no single prompting strategy generalized across models, and lower decoding temperatures improved stability across tasks. These results underscore that prompt engineering remains model-specific and empirically driven rather than transferable across architectures.

### Clinical implications

Beyond benchmarking, this study demonstrates the clinical utility of LLM-based phenotyping for scalable extraction of patient-reported self-management behaviors in rheumatology and potentially other specialties. The pipeline identified decade-long cannabis trends, with pain as the predominant reason for use, followed by sleep and nausea. Pain scores shifted over time: users reported higher scores earlier in the decade, but the gap narrowed and reversed in recent years, consistent with mixed evidence on cannabis efficacy and substantial individual variability in pain management(2,4). Although causal inference is not possible, these text-derived phenotypes can identify evolving real-world patterns for future mechanistic or prospective study.

### Limitations

Several limitations should be considered. First, clinical documentation of cannabis use is not standardized and likely underrepresents true prevalence due to variable disclosure and clinician documentation practices. Second, snippet-level extraction may not capture full narrative context, particularly when motivations are distributed across longer text spans. Third, substantial class imbalance, especially for reason-of-use categories, may influence performance metrics and minority class sensitivity. Fourth, patient-level classification based on the presence of at least one “current use” snippet may introduce misclassification if isolated mentions are erroneous. Finally, the study was conducted within a single academic health system, and external validation across institutions is needed to assess generalizability.

### Conclusion

This study shows when and how LLMs add value for clinical information extraction. Domain-adapted transformers may be more efficient and robust for structured classification, whereas large generative LLMs may help with context-rich, semantically variable extraction. Rather than supporting universal LLM superiority, our results favor task-specific deployment that balances performance, computational cost, and generalizability. The benchmarking and application framework can support clinical phenotyping workflows and real-world research on substance use and symptom management in chronic disease populations.

## Data Availability

Data access is restricted in accordance with institutional privacy regulations and ethical approvals.

https://github.com/yiyuwang/llm_ard_scripts

## Acknowledgement

YW is supported by the National Institute on Drug Abuse T32DA035165. TF is supported by the National Institute of Arthritis and Musculoskeletal and Skin Diseases K01AR079039. We used OpenAI’s ChatGPT to assist with language editing and improving the readability of the manuscript.

## Data and Code Availability

Data access is restricted in accordance with institutional privacy regulations and ethical approvals. Analysis scripts are available at https://github.com/yiyuwang/llm_ard_scripts

## Author Contributions

YW and SB are joint first authors. YW performed the data analysis, with NL provided analytical support. YW, SB, TF designed the study. TF, AA, and AH provided clinical guidance and notes annotation. TF managed data acquisition. YW and SB wrote the manuscript with all authors contributing to reviewing.

## Ethics Approval

The study was approved by the Stanford University Institutional Review Board (IRB #53750) with a waiver of informed consent for the secondary use of de-identified data. No patients or members of the public were involved in the design, conduct, reporting, or dissemination of this research.

