## Supplemental Materials for "Extracting patient reported cannabis use and reasons for use from electronic health records: a benchmarking study of large language models"

**Running Title:** Cannabis Use LLM Methods

**Manuscript Type:** Original Research

Supplementary Materials

Figure S1. Data Flow.

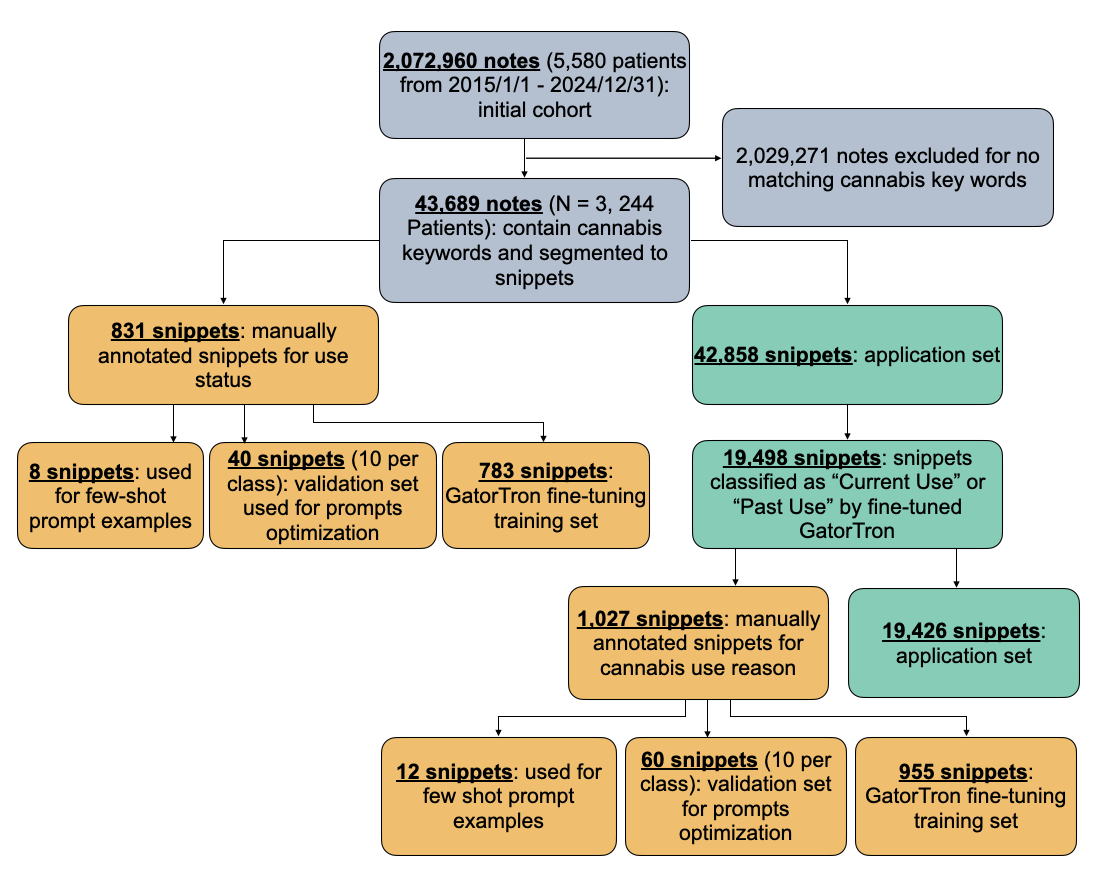

Figure S2. Model Performance Comparison across Temperature and Strategy: Use Status

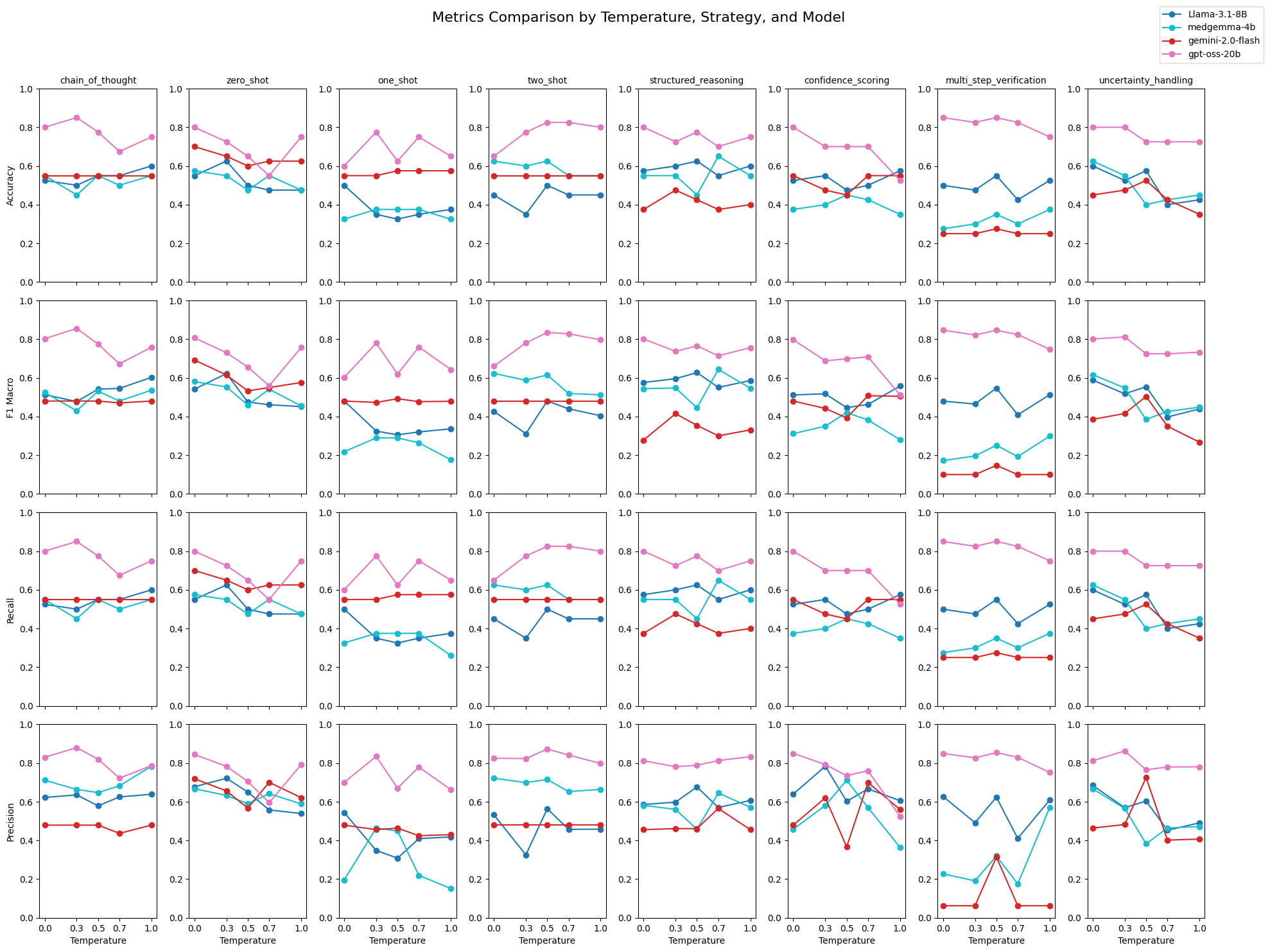

Figure S3. Model Performance Comparison across Temperature and Strategy: Use Reason
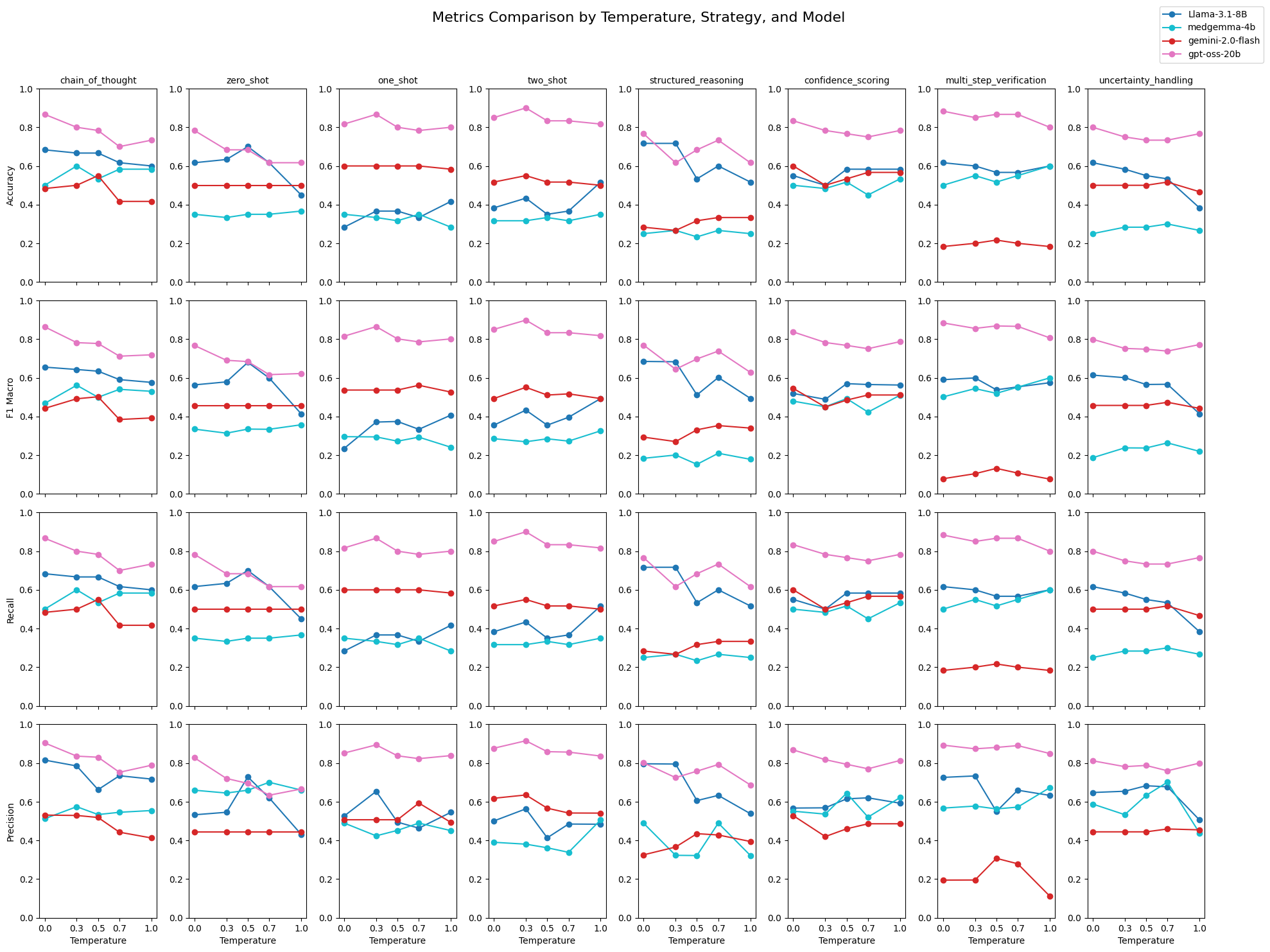

Table S1. Prompts used for use status

| Chain of thought | You are an experienced clinical assistant in substance use.  Your task is to identify cannabis use from clinical notes with high accuracy.  You will read the following clinical snippet and THINK STEP BY STEP about whether it indicates cannabis use from the patient.  First, explain your reasoning in one sentence. Then classify the snippet as:  1 = Not A True Mention  2 = Denial of Use  3 = Positive Past Use  4 = Positive Current Use    Snippet:  {text}  After your reasoning, respond with a JSON object in this exact format:  {{"classification": <number between 1-4>}} |
| --- | --- |
| Zero shot | You are an experienced clinical assistant in substance use.  Your task is to identify cannabis use from clinical notes with high accuracy.  Review this snippet as you would in your professional practice:  {text}  Based on your expertise, classify this mention:  1 = Not A True Mention  2 = Denial of Use  3 = Positive Past Use  4 = Positive Current Use  Respond with a JSON object in this exact format:  {{"classification": <number between 1-4>}} |
| One shot* | You are an experienced clinical assistant in substance use.  Your task is to identify cannabis use from clinical notes with high accuracy.  Here are examples for your reference:  Example 1: " "  Classification: 1 (Not A True Mention)  Example 2: " "  Classification: 2 (Denial of Use)  Example 3: " "  Classification: 3 (Positive Past Use)  Example 4: " "  Classification: 4 (Positive Current Use)  Now classify this snippet:  {text}  Respond with a JSON object in this exact format:  {{"classification": <number between 1-4>}} |
| Two-shot* | You are an experienced clinical assistant in substance use.  Your task is to identify cannabis use from clinical notes with high accuracy.  Here are examples for your reference:  Example: " "  Classification: 1 (Not A True Mention)  Example: " "  Classification: 1 (Not A True Mention)  Example: " "  Classification: 2 (Denial of Use)  Example: " "  Classification: 2 (Denial of Use)  Example: " "  Classification: 3 (Positive Past Use)  Example: " "  Classification: 3 (Positive Past Use)  Example: " "  Classification: 4 (Positive Current Use)  Example: " "  Classification: 4 (Positive Current Use)  Now classify this snippet:  {text}  Respond with a JSON object in this exact format:  {{"classification": <number between 1-4>}} |
| Structured reasoning | You are an experienced clinical assistant in substance use.  Your task is to identify cannabis use from clinical notes with high accuracy using the following framework:  ANALYSIS:  1. Context identification: [Is this about the patient, family history, or general discussion?]  2. Temporal markers: [Look for words indicating past, present, or future]  3. Negation detection: [Check for denial, "no", "denies", etc.]  4. Certainty assessment: [Is the statement definitive or uncertain?]  CLASSIFICATION:  1 = Not A True Mention  2 = Denial of Use  3 = Positive Past Use  4 = Positive Current Use  SNIPPET: {text}  After your analysis, respond with a JSON object in this exact format:  {{"classification": <number between 1-4>}} |
| Confidence scoring | You are an experienced clinical assistant in substance use.  Your task is to identify cannabis use from clinical notes with high accuracy.  Classify this clinical snippet for cannabis use and provide your confidence level:  Classification:  1 = Not A True Mention  2 = Denial of Use  3 = Positive Past Use  4 = Positive Current Use  Snippet: {text}  First provide your confidence level and reasoning, then respond with a JSON object containing your classification:  {{"classification": <number between 1-4>, "confidence": "<Low/Medium/High>", "reasoning": "<brief explanation>"}} |
| Multi-step verification | You are an experienced clinical assistant in substance use.  Your task is to identify cannabis use from clinical notes with high accuracy using the following verification steps:  Step 1: Does this snippet mention cannabis/marijuana/THC or related terms? (Yes/No)  Step 2: If yes, is this referring to the patient specifically? (Yes/No/Unclear)  Step 3: What is the temporal context?  1 = Not A True Mention  2 = Denial of Use  3 = Positive Past Use  4 = Positive Current Use    Step 4: Final classification for snippet: "{text}"  After your reasoning, respond with a JSON object in this exact format:  {{"classification": <number between 1-4>}} |
| Uncertainty Handling | You are an experienced clinical assistant in substance use.  Your task is to identify cannabis use from clinical notes with high accuracy.  Ambiguity is common in clinical notes. If you encounter uncertainty, use classification 1.  Snippet: {text}  Analysis:  - Clear indicators present: [Yes/No]  - Ambiguous elements: [List any unclear aspects]  - Missing context: [What information would help?]  If certain, provide classification 1-4:  1 = Not A True Mention  2 = Denial of Use  3 = Positive Past Use  4 = Positive Current Use  After your analysis, respond with a JSON object in this exact format:  {{"classification": <number between 1-4>, "certainty": "<High/Medium/Low>"}} |

* example snippets were removed in the table for patient privacy

Table S2. Prompts for use reasons

| Chain of thought | You are an experienced clinical assistant in substance use.  Your task is to identify the primary reason for cannabis use from clinical notes with high accuracy.  You will read the following clinical snippet and THINK STEP BY STEP about the reason for the patient's cannabis use.  First, explain your reasoning in detail. Then classify the primary reason for cannabis use as:  1 = Current Use For Pain  2 = Current Use For Nausea  3 = Current Use For Sleep  4 = Current Use For Relaxation / Stress / Anxiety  5 = Current Use For Appetite  6 = Not Applicable / Unknown  Snippet:  {text}  After your reasoning, respond with a JSON object in this exact format:  {{"classification": <number between 1-6>}} |
| --- | --- |
| Zero shot | You are an experienced clinical assistant in substance use.  Your task is to identify the primary reason for cannabis use from clinical notes with high accuracy.  Review this snippet as you would in your professional practice:  {text}  Based on your expertise, classify the primary reason for cannabis use:  1 = Current Use For Pain  2 = Current Use For Nausea  3 = Current Use For Sleep  4 = Current Use For Relaxation / Stress / Anxiety  5 = Current Use For Appetite  6 = Not Applicable / Unknown  Respond with a JSON object in this exact format:  {{"classification": <number between 1-6>}} |
| One shot* | You are an experienced clinical assistant in substance use.  Your task is to identify the primary reason for cannabis use from clinical notes with high accuracy.  Here are some examples for each classification:  Example 1: " "  Classification: 1 (Current Use For Pain)  Example 2: " "  Classification: 2 (Current Use For Nausea)  Example 3: " "  Classification: 3 (Current Use For Sleep)  Example 4: " "  Classification: 4 (Current Use For Relaxation / Stress / Anxiety)  Example 5: " "  Classification: 5 (Current Use For Appetite)  Example 6: " "  Classification: 6 ( Not Applicable / Unknown )  Now classify this snippet:  {text}  Respond with a JSON object in this exact format:  {{"classification": <number between 1-6>}} |
| Two-shot* | You are an experienced clinical assistant in substance use.  Your task is to identify the primary reason for cannabis use from clinical notes with high accuracy.  Here are some examples for each classification:  Example: " "  Classification: 1 (Current Use For Pain)  Example: " "  Classification: 1 (Current Use For Pain)  Example: " "  Classification: 2 (Current Use For Nausea)  Example: " "  Classification: 2 (Current Use For Nausea)  Example: " "  Classification: 3 (Current Use For Sleep)  Example: " "  Classification: 3 (Current Use For Sleep)  Example: " "  Classification: 4 (Current Use For Relaxation / Stress / Anxiety)  Example: " "  Classification: 4 (Current Use For Relaxation / Stress / Anxiety)  Example: " "  Classification: 5 (Current Use For Appetite)  Example: " "  Classification: 5 (Current Use For Appetite)  Example: " l"  Classification: 6 ( Not Applicable / Unknown )  Example: " "  Classification: 6 ( Not Applicable / Unknown )  Now classify this snippet:  {text}  Respond with a JSON object in this exact format:  {{"classification": <number between 1-6>}} |
| Structured reasoning | You are an experienced clinical assistant in substance use.  Your task is to identify the primary reason for cannabis use from clinical notes with high accuracy.  Analyze this clinical snippet for the reason behind cannabis use using the following framework:  ANALYSIS:  1. Presence of cannabis use: [Is cannabis use mentioned?]  2. Primary symptoms addressed: [Pain, nausea, sleep issues, anxiety, appetite, etc.]  3. Contextual indicators: [Words that connect the use to specific symptoms]  4. Alternative purposes: [Any other reasons mentioned]  CLASSIFICATION:  1 = Current Use For Pain  2 = Current Use For Nausea  3 = Current Use For Sleep  4 = Current Use For Relaxation / Stress / Anxiety  5 = Current Use For Appetite  6 = Not Applicable / Unknown  SNIPPET: {text}  After your analysis, respond with a JSON object in this exact format:  {{"classification": <number between 1-6>}} |
| Confidence scoring | You are an experienced clinical assistant in substance use.  Your task is to identify the primary reason for cannabis use from clinical notes with high accuracy.  Classify the primary reason for cannabis use in this clinical snippet and provide your confidence level:  Classification:  1 = Current Use For Pain  2 = Current Use For Nausea  3 = Current Use For Sleep  4 = Current Use For Relaxation / Stress / Anxiety  5 = Current Use For Appetite  6 = Not Applicable / Unknown  Snippet: {text}  First provide your confidence level and reasoning, then respond with a JSON object containing your classification:  {{"classification": <number between 1-7>, "confidence": "<Low/Medium/High>", "reasoning": "<brief explanation>"}} |
| Multi step verification | You are an experienced clinical assistant in substance use.  Your task is to identify the primary reason for cannabis use from clinical notes with high accuracy.  Step 1: Does this snippet mention cannabis/marijuana/THC use? (Yes/No)  Step 2: If yes, what appears to be the primary purpose of use?  - Is pain management mentioned? (Yes/No)  - Is nausea management mentioned? (Yes/No)  - Is sleep improvement mentioned? (Yes/No)  - Is relaxation, stress or anxiety management mentioned? (Yes/No)  - Is appetite stimulation mentioned? (Yes/No)  - Is another reason mentioned or is the reason unclear? (Yes/No)  Step 3: Final classification for snippet: "{text}"  1 = Current Use For Pain  2 = Current Use For Nausea  3 = Current Use For Sleep  4 = Current Use For Relaxation / Stress / Anxiety  5 = Current Use For Appetite  6 = Not Applicable / Unknown  After your reasoning, respond with a JSON object in this exact format:  {{"classification": <number between 1-6>}} |
| Uncertainty Handling | You are an experienced clinical assistant in substance use.  Your task is to identify the primary reason for cannabis use from clinical notes with high accuracy. If multiple reasons are mentioned, classify by the primary/most emphasized reason.  Ambiguity is common in clinical notes. If you encounter uncertainty, use classification 6.  Snippet: {text}  Analysis:  - Cannabis use mentioned: [Yes/No]  - Indicators of reason: [List any mentioned reasons]  - Primary reason (if multiple): [Which reason seems most important?]  - Ambiguous elements: [List any unclear aspects]  Classification options:  1 = Current Use For Pain  2 = Current Use For Nausea  3 = Current Use For Sleep  4 = Current Use For Relaxation / Stress / Anxiety  5 = Current Use For Appetite  6 = Not Applicable / Unknown  After your analysis, respond with a JSON object in this exact format:  {{"classification": <number between 1-6>, "certainty": "<High/Medium/Low>"}} |

* example snippets were removed in the table for patient privacy

Table S3. Final Hyperparameters for Fine-Tuned GatorTron

|  | Learning rate | Dropout rate | Maximum token length | Full fine-tune or Linear Probe |
| --- | --- | --- | --- | --- |
| Cannabis Use Status GatorTron | 5.29 × 10⁻^5^ | 0.15 | 192 | Full fine-tune |
| Cannabis Use Reason GatorTron | 1.16 × 10⁻⁴ | 0.15 | 128 | Full fine-tune |
